# The demand of cancer patients to anticipate the management of undernutrition: is a nutrition prehabilitation before treatment conceivable as supportive care to improve quality of life? Argument for the NEHOTEL® concept

**DOI:** 10.1101/2024.02.13.24302761

**Authors:** Pouillart Philippe, Dépeint Flore, Soula Isabelle, Raynard Bruno

## Abstract

**Purpose:** Our translational research team in nutrition and cancer works to identify changes in eating and cooking behaviors since the diagnosis. Educational tools have been standardized as supportive care workshops in culinary practice and nutrition to enhance quality of life with regards to symptoms associated with treatment. Ongoing work with a cluster of expert patients led to the conclusion that such tools are essential but may be addressed too late, once symptoms are present and undernutrition is set. We thus investigated the concept of an early nutritional and multimodal prehabilitation program to improve quality of life.

**Methods:** Over a period of one year, 10 patients, together with researchers and caregivers co-constructed the NEHOTEL educational program. The relevance of this program was then confronted to the practical, medical and ethical points of view.

**Results:** An innovative multimodal supportive care 5-days course has been validated with our panel of expert patients. In this setting, new cancer patients will be invited to a non-medical hotel facility specifically designed for this project.

**Conclusions:** In the synopsis of the cancer care path as currently conceived in France, the early positioning of this intervention, which formalizes the need for unmet supportive care of patients in nutrition, raises questions about its medico-technical feasibility and the chances of visualizing a benefit on the quality of life. NEHOTEL® concept design is the outcome of this translational work, supported by an ongoing clinical feasibility study.

## Introduction

Patient-centered care is now recognized as a benchmark of quality care for people who are affected by chronic conditions such as cancer. Effective, high quality cancer care is viewed as involving more than just the delivery of anti-cancer therapy and, increasingly, cancer service providers are required to address patients’ supportive care needs in nutrition [1]. Indeed, cancer patients are at high risk of losing vital body energy resources due to the pathology hypermetabolism. Cumulated with treatments side effects, this situation results in systemic inflammatory response syndrome (SIRS) driving global catabolism, malnutrition and sarcopenia, impaired quality of life and worse clinical outcome [2]. Malnutrition is accepted as both a cause and a consequence of treatment side effects [3]. Weight loss is frequent, being among the first symptoms in more than 50% of patients and occurring in over 70% during the later course of the disease. Although the stage and type of cancer and the response to chemotherapy are the most important prognostic factors, many studies have reported that patients who maintained weight presented better prognosis than those who did not [4]. Nutritional status has a prognostic value and optimal supportive care in nutrition is required to allow patients to tolerate aggressive or long-term anticancer treatments, to maintain an adequate quality of life and to ensure the course of treatments is completed according to the prescription. We are among those who believe that nutritional intervention should be engaged as early as 5% of body weight loss and that we need simple low-cost methods such as Visual/Verbal Analogue Scale of food ingesta (ingesta-VVAS) to be used during the screening carried out for every inpatient [5, 6].

According to the reviewed data and guidelines, nutritional recommendations should be systematic as adjuvant to any treatment and should be included in the multidisciplinary approach mandatory in oncology [2, 4, 7, 8]. This will allow for more adequate and efficient results in patients at risk of malnutrition. Recommendations need to address and focus on all physical, psychological and social problems interfering with food intake, digestion and anabolism to maintaining adequate body resources and functions [2]. Nutrition counseling is a major therapeutic option as a non-pharmaceutical intervention solution (NPIS). Textured and enriched foods should be offered to patients with a functional intestinal tract whenever possible before proposing oral nutritional supplements, then enteral or parenteral nutrition [9]. Cooking practice is a simple, natural and familiar nutritional intervention and early education is of major importance in oncology, thus being a key factor for successful treatment and recovery (Figure 1).

**Figure 1.**
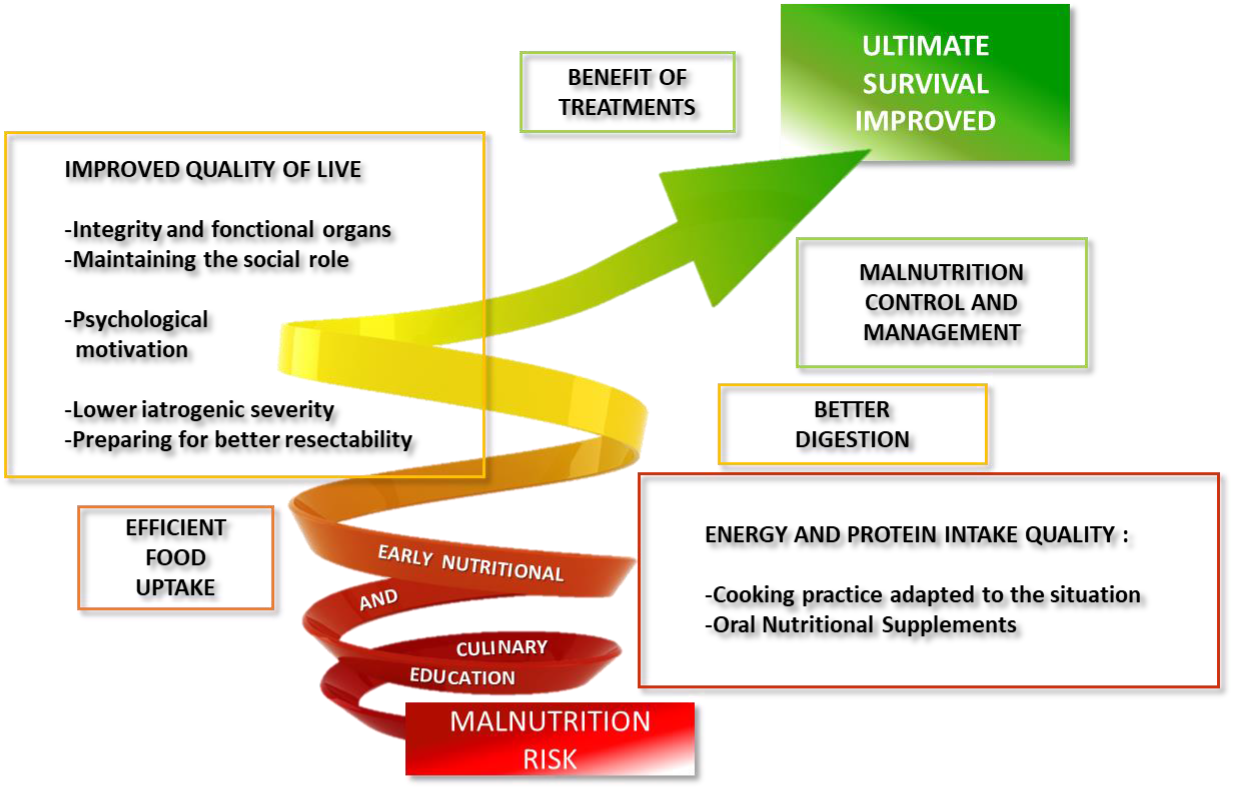
Nutrition counselling and early adapted cooking practices are essential choices to prevent or manage malnutrition in cancer patients with a functional intestinal tract.

Cancer patients tend to be obsessed with food control, and thus they could actively participate in the cancer treatment process by implementing appropriate diet modifications. But such counseling needs to be validated and promoted by caregivers and scientific communities. Even if we are inspired by patients we need to stay driven by science. It is why our nutrition and cancer translational program NEODIA is coordinated by a staff associating supportive care educators, dieticians, a chef, a pharmacognosist, as well as oncologists and other associated medical staff.

In a survey initiated in 2010, we interviewed 197 patients who had been under treatment for 6 to 12 months for a cancer in active phase (all situations and therapeutic protocols). A series of 125-questions was addressed, focusing on symptoms that disrupt daily life and their impact on eating and cooking habits. Patients were also asked about their level of knowledge on nutrition and possible access to dietary information during their care pathway and who provided it. Cross-sectional observational studies are typically limited to extracting trends in food consumption and habits. This is even more complex here since patient profiles are transversal to different cancers, various therapeutic protocols, length of treatment and evolutionary symptoms. However, the main objective was to identify the adaptations that the patients had adopted to alleviate the symptoms they experienced. In the vast majority of cases, whatever the etiology of side effects which may be related to different treatments, nutritional solutions were the same as long as the symptom was temporary and of moderate amplitude.

In agreement with literature [10], our survey shows that (1) one in three patients had deserted their kitchen and needed help to prepare meals. Three out of 4 patients were concerned with fatigue, asthenia and nausea, and were not able to resume cooking and meal preparation immediately after treatment. Patients’ parents or partner were the main source of help in the kitchen; (2) One third considered the meal to be an unpleasant moment, half of them dreading the meal time; (3) 54.9% of patients identified breakfast as their favorite meal; and (4) Two thirds of patients considered that food has an effect on the body during treatments. As a consequence, patients reported that they (or partners) had adapted ingredients in recipes or modified cooking practices, to favor some foods and beverages rather than avoidance of specific food items. Patients also used a wide range of aromatic plants and mild spices as small and repeated doses throughout the day in dishes as well as infusions to relieve symptoms [11](Table 1).

**Table 1.**
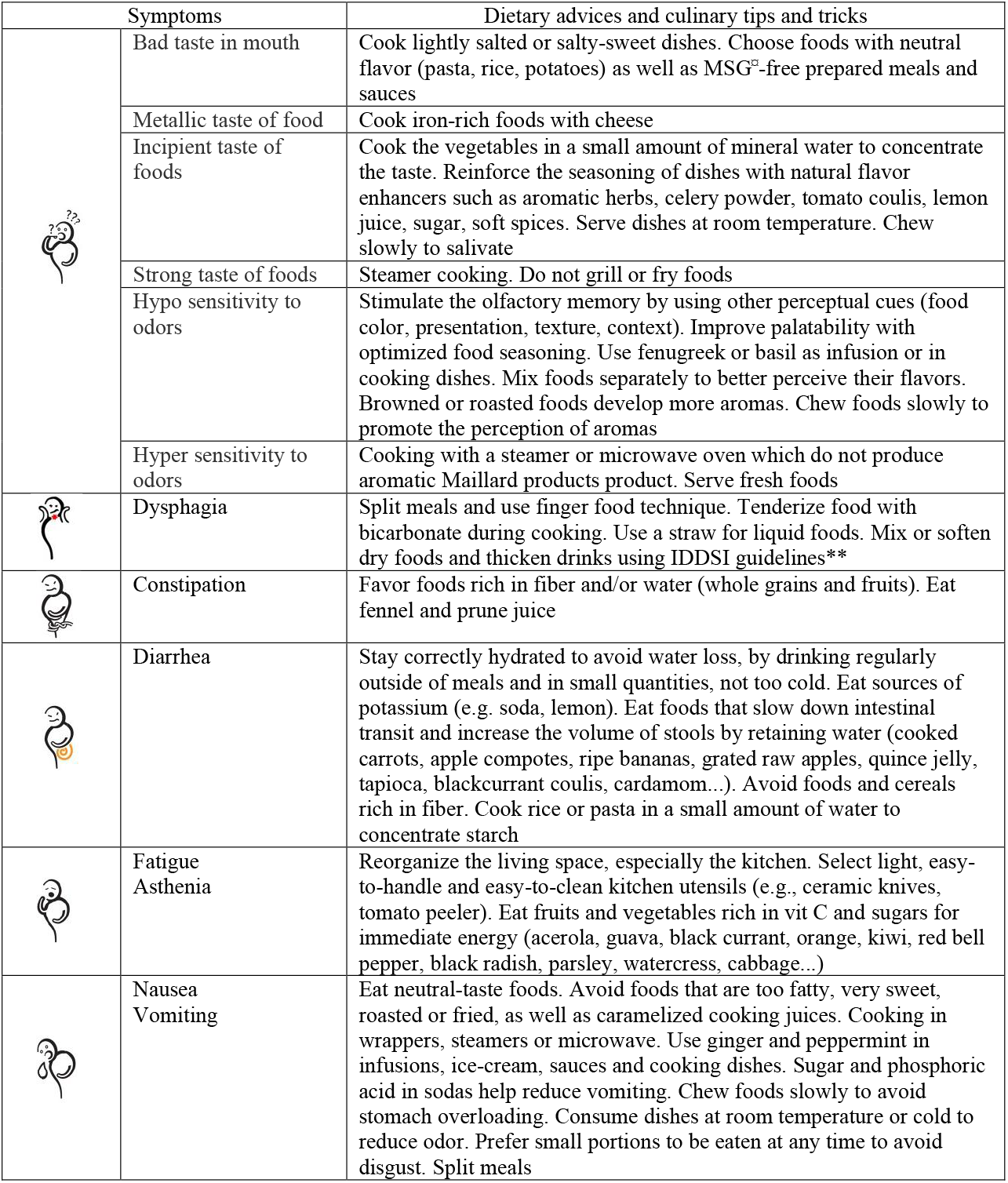
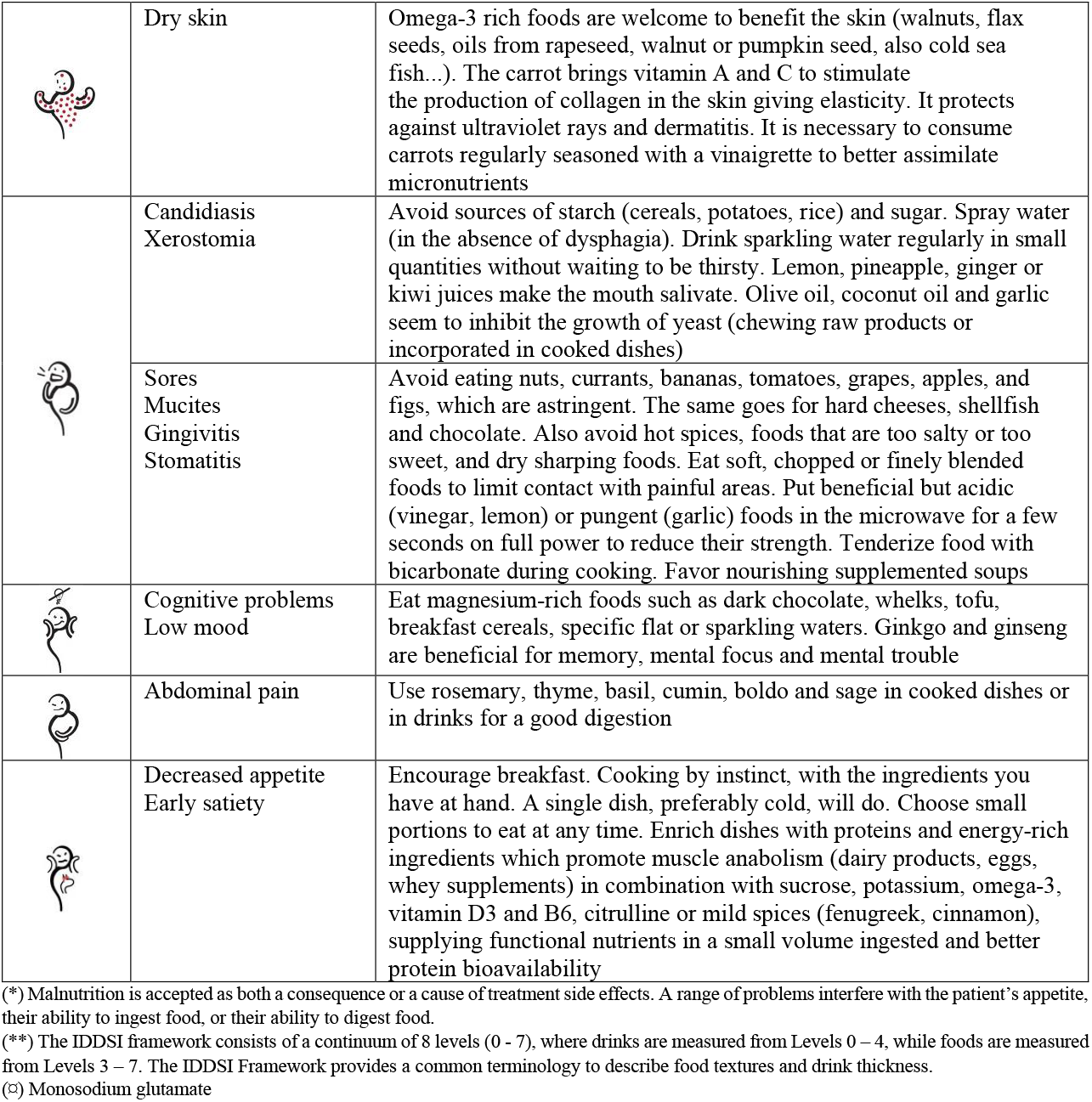
Responses to common direct or undirect causes for a poor nutritional status in cancer patients*: synthesis of dietary advices and culinary tips and tricks from the NEODIA translational group of research to lower symptoms.

In a follow-up study, 40 cancer patients from the initial survey group were included in hedonics tests. As compared to a control group (n=40), they seemed to no longer seek pleasure in individual foods but looked on dishes evoking conviviality, sharing and naturality [12]. As previously described, food hedonics are broader than the eating and drinking experience [13]. They may include a good palatability but also a psychological determination of the extent to which diet provides emotion of pleasure or displeasure. Furthermore, we need to take in account that the mental perception shift suggesting alteration of food preferences, as observed with sugar consumption is polluted by fake news on social networks “sugar feeds cancer” [14].

In another study including patients in a week-long hospital stay, criteria for choosing catered food ranked as follow: flavor was the main criterion, followed by odors, dish presentation, serving temperature, with quantity of food ranking last [12]. Similar criteria were evaluated for acceptability of dishes proposed by the hospital catering service [15]. Results were in line with a national French study [10].

As a result, and since 2017, we transferred this knowledge into hospital chefs’ training programs considering that hospital catering is a continuation of medical prescription [16]. Tips to manage proteins and energy density, as well as modified food textures and finger food are taught in line with guidelines of the international dysphagia diet standardization initiative (IDDSI) [17]. Enriching dishes and beverages with efficient proteins such as whey can promote muscle anabolism and prevent sarcopenia. If consumed in combination with carbohydrates, potassium, omega-3, vitamin D3 and B6, citrulline, as well as mild spices, the meal supplies essential and functional nutrients in small portions and improves protein bioavailability [11, 12, 18]. These nutritional recommendations have been recently updated and biochemical pathways well documented [2, 4]. If we consider culinary uses of mild spices and aromatic plants to alleviate symptoms, and recipes associating a variety of valuable macro- and micronutrients able to promote muscle protein anabolism and reducing inflammation [19], and by conforming to circadian rhythm’s favoring protein accretion in the first part of the dietary day for a better physical functioning [20] (ie. breakfast “of a king”, lunch “of a prince”, dinner “of a poor”), the interest of the Mediterranean diet during cancer is emerging in nutritional counseling as supportive care to patients. We have also integrated this knowledge in education programs for patients and caregivers [21].

Table 1 is a global synthesis of 10-years of studies at UniLaSalle into providing the right dietary and cooking advice according to the symptoms. This work was also highlighted by a book, a large number of presentations in supportive care conferences and various publications in French published in professional and technical journals to directly reach both hospital services, including caregivers and catering, and patients’ associations as well as loved ones (Table 2). We also shared our results to a wider audience by launching in 2015 the culinary web site https://vite-fait-bienfaits.fr (ie: “quickly and well done with health benefits”) in which the editorial content, recipes and ergonomics were co-constructed with expert patients.

**Table 2.**
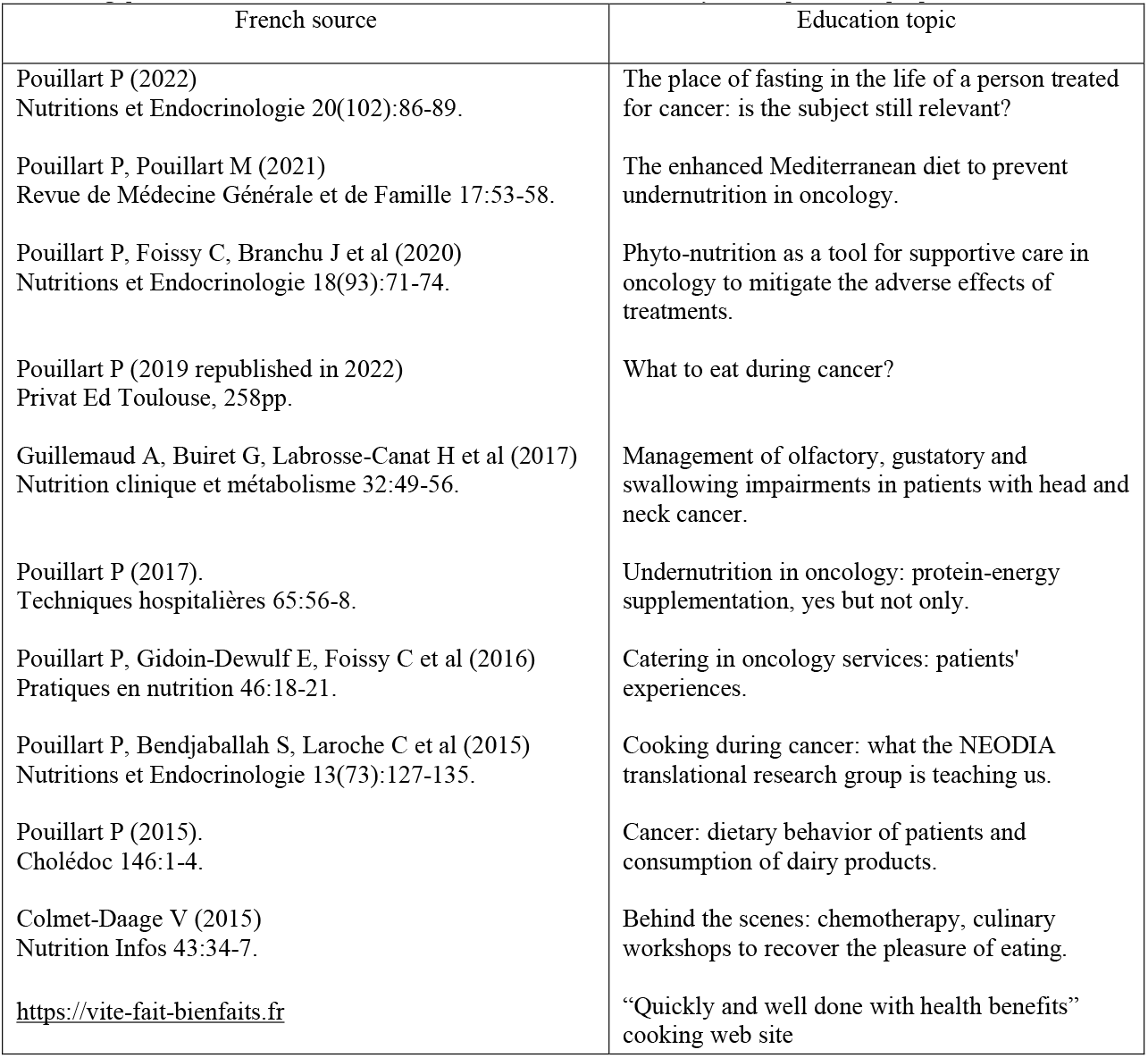
Expertise transfer from the NEODIA translational group of research to French hospital services including caregivers and catering, patients’ associations and as well as home loved ones to directly reach operational people.

Since 2015, our patient-centered data have been regularly compared with evidence-based guidance of consensus reports and then integrated into a supportive care standardized culinary workshop for both hospitals and patients’ associations settings. This “vite-fait-bienfaits” model of culinary workshop is 2.5 hours long and includes cooking practice and tasting as well as informal exchanges in groups of 6 to 8 patients. It is currently in place in 20 structures in France, Belgium and Switzerland [22]. During the Covid-19 pandemic, frequency of face-to-face workshops was disrupted. However, visits to the vite-fait-bienfaits.fr website increased by 20% to reach 180,000 connections a year. The pandemic situation led us to design supportive care sessions in the form of live nutrition cookinars dedicated to patients and their families and validated by practitioners (publication in progress). The continuity of supportive care workshops in nutrition is important to accompany the medical course according to the needs of the patients, and the first distance learning sessions were very well received by patients and loved one.

However, our 10 expert patients reminded us that they experienced early symptoms precursor of undernutrition without having solid knowledge to face them, even though their pathological situation was known to be “at risk” (digestive or oesogastric cancers and aggressive therapeutic protocols). It is why they proposed to discuss the feasibility of early supportive care prehabilitation in nutrition for newly diagnosed people. In this publication we describe our collective approach and the resulting program.

## Methods

### Expert patient panel

Initially, we included 10 of the respondents to the 2010 survey. Selection criteria included successful application of culinary and dietary tips and tricks to minimize symptoms, home-based living and implication in daily preparation of meals, as well as willingness to participate in further work with researchers at UniLaSalle. The group was trained in research methodology. All 10 initial expert patients were women. Whenever a patient left the group, and whatever the reason, she was replaced by a new patient with the same profile addressed by local patients’ associations. The group that worked on the NEHOTEL^®^ project was composed of 3 women with cancer recurrence from the 2010 survey and 7 women integrated more recently (53 to 66 years old).

In addition to development of new recipes, expert patients participated in design of the names and logos associated with the various works of the translational team. These include (1) “NEODIA” (ie: NEO: “cancer” / DIA: “journey through the care pathway”) and (2) “vite-fait-bienfaits” (ie: “quickly and well done with health benefits”) in the early years of the group, as well as (3) NEHOTEL^®^ (ie: NEO: “cancer” / HOTEL: “the concept to welcome cancer patients in a supportive care program focused on nutrition”) and (4) NEHOTEL Resort^®^ (ie: NEO: “cancer” / HOTEL Resort: “a hotel-restaurant facility specially dedicated to receive patient included in NEHOTEL^®^ as an all-inclusive service”) for the work presented herein.

### Development of the NEHOTEL^®^ concept

Expert patients were invited to participate in creative workshops at different times for a total of 10 days. During the first 4 sessions, brainstorming exercises with two supportive care professionals allowed expert patients to freely verbalize and formalize the need for unmet supportive care programs. A group of supportive care professionals (dietetics, socio-aesthetic, adapted physical activity, sophrology, psychology) then analyzed the raw demand to create a draft of the educative content. This was followed by a 4-days session with expert patients to discuss and adjust the final educative synopsis. The specificities of the hospitality facility or venue that would host such a program was then addressed. The relevance and potential feasibility of the program was studied and compared to the literature by the medical staff of Beauvais Hospital. A final proposition from medical professionals was assessed by expert patients and supportive care professionals during a 2-days workshop.

### Clinical validation of the NEHOTEL^®^ concept

Before the program can apply for therapeutic education of patient (TEP) label, it is important to evaluate its feasibility, efficacity and economic impact. With this objective, a clinical trial protocol (2021-A00901-40) has been approved by ethics committee “Ile-de-France III” on June 6^th^ 2022. The protocol has been designed with oncology and supportive care professionals together with a clinical research associate. Quality assurance engineers have coordinated the development of NEHOTEL Resort^®^. The study is also followed by a review committee including 5 experts in the fields of oncology, supportive care, nutrition and patients’ associations.

The one-arm intervention trial will assess adherence to the program, efficacy on quality of life and medical outcome, as well as the technico-economic feasibility of the concept. A total of 60 participants will be addressed from 5 hospitals in Hauts-de-France. Undernutrition is integral to the rate of cancer spread, particularly for cancers with a prevalence of undernutrition greater than 45% [9]. Thus, we decided to include patients with head and neck, bronco-pulmonary, pancreas, oeso-gastric and duodenal cancers, excluding ovarian, kidney or bladder cancers which present a late risk of undernutrition and need a different supportive care approach in nutrition. NEHOTEL^®^ clinical trial inclusion and non-inclusion criteria as well as clinical trial outcomes are presented respectively in Tables 3 and 4. Undernutrition is a predictive factor of the prognosis at 8 months in the targeted cancers [23]. NEHOTEL^®^ participants will be remotely followed for 8 months after the program using a dedicated digital application for dietetics (DIETIS^®^, ALANTAYA Co., France). The study does not modify in any way the therapeutic protocol proposed by the medical multidisciplinary commission of the addressing hospitals and patients will remain under their care.

**Table 3.** NEHOTEL^®^ clinical trial inclusion (I) and non-inclusion (NI) criteria.

|  |
| --- |
| Clinical trial inclusion (I) criteria<br>I1: Head & neck, pancreatic, oesogastric, duodenal or broncho-pulmonary<br>I2: Patient who have not begun their treatments or patient in the beginning of their treatments, less than or equal to two sessions of chemotherapy or radiotherapy<br>I3: Male or female<br>I4: From 18 to 70 years old<br>I5: Patient living in a private home (personal or family)<br>I6: Prognosis greater than 12 months<br>I7: Decision taken by multidisciplinary consultation staff for a curative treatment<br>I8: WHO Score Performance Status $\leq 2$<br>I9: Patient undernourished at the advertisement codes E44.1 "Mild protein-energy malnutrition" or E44.0 "Moderate protein-energy malnutrition" or<br>I10: Patient not undernourished at the time of the announcement, but going to receive a treatment or combination of treatments for curative purposes, whose therapeutic sequences are known to induce nutritional complications inducing a risk of stopping treatment<br>I11: Per os feeding<br>I12: Internet access and reachable by phone<br>I13: Patient affiliated to the social security system |
| I14: Patient information and signature of informed consent<br>I15: Patient accompanied by a family member or not |
| Clinical trial non-inclusion (NI) criteria<br>NI1: Patient with severe malnutrition corresponding to at least one of the criteria below, taking in account that a single criterion of severe undernutrition takes precedence over one or more criteria of moderate undernutrition:<br>(i) Body Mass Index (BMI) < 17 kg/m<sup>2</sup><br>(ii) Weight loss ≥ 10% in 1 month or ≥ 15% in 6 months or ≥ 15% of the usual weight before the onset of the disease<br>(iii) Albuminemia ≤ 30 g/L<br>NI2: Treatment for curative purposes not applicable<br>NI3: Comorbidities that do not allow participation in the prehabilitation course (patient presenting at least one of the criteria below):<br>(i) Weight greater than 130kgs (limit of resistance of the NEHOTEL® beds)<br>(ii) Sensory deficits: visual, auditory, olfactory, gustatory (not allowing to follow the educational workshops)<br>(iii) Cognitive deficit (reading, writing, counting)<br>(iiii) Physical (ability to move around and participate in activities)<br>(iiiii) Patient at risk of alcohol withdrawal<br>NI4: TNM (Tumor Node Metastasis) coupled with a deteriorated general condition of the patient at the time of the announcement; prediction of highly mutilating surgery involving an inability to eat through the mouth; metastatic stage.<br>NI5: Patient institutionalized and/or not responsible for his or her diet<br>NI6: Patient requiring parenteral or artificial enteral nutrition (feeding tube, nasogastric tube, gastrostomy or feeding jejunostomy) |

**Table 4.**
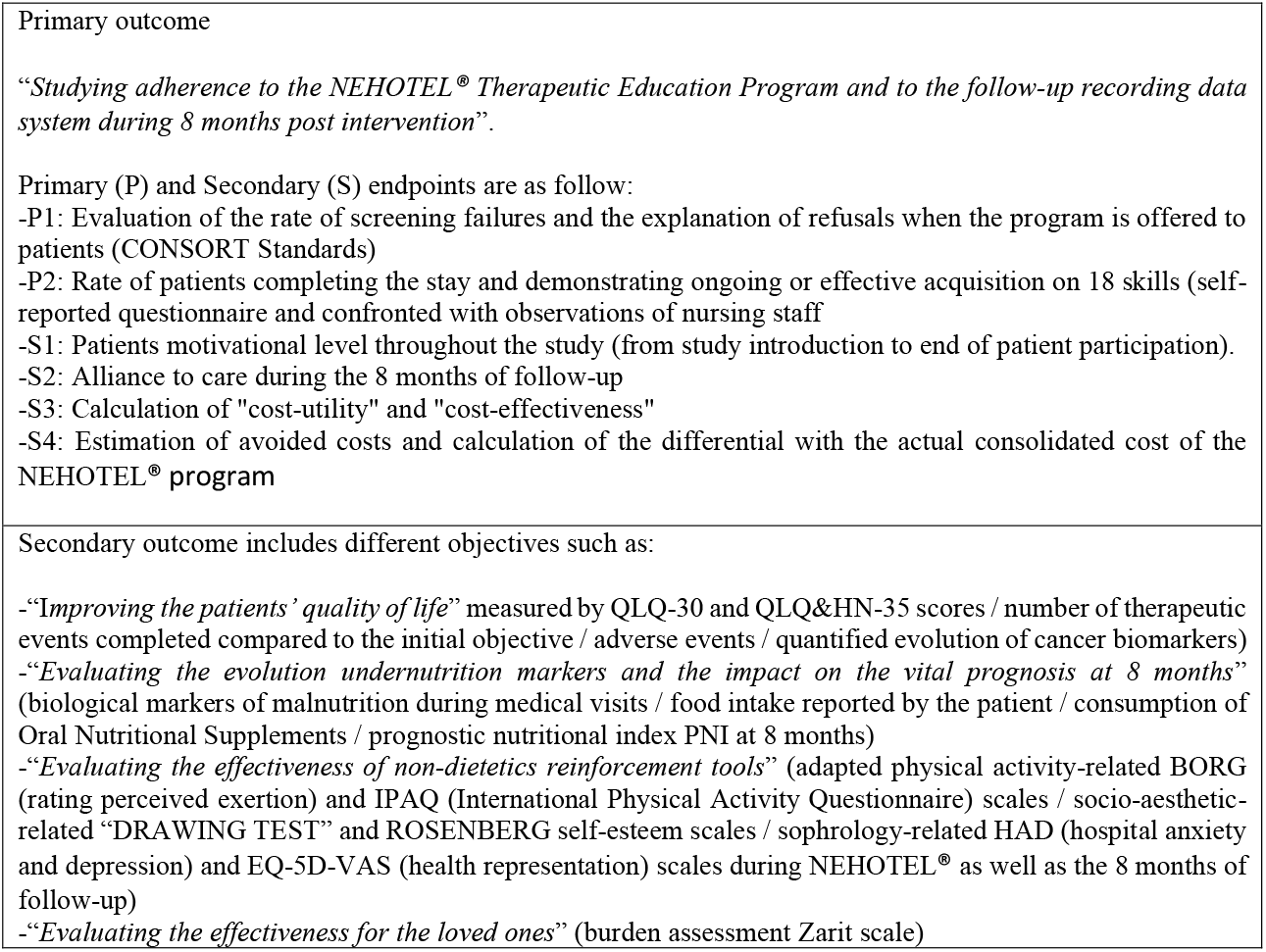
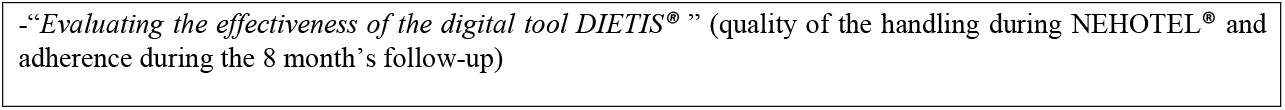
NEHOTEL^®^ clinical trial outcomes.

|  |
| --- |
| Primary outcome<br>*“Studying adherence to the NEHOTEL® Therapeutic Education Program and to the follow-up recording data system during 8 months post intervention”.*<br>Primary (P) and Secondary (S) endpoints are as follow:<br>-P1: Evaluation of the rate of screening failures and the explanation of refusals when the program is offered to patients (CONSORT Standards)<br>-P2: Rate of patients completing the stay and demonstrating ongoing or effective acquisition on 18 skills (self-reported questionnaire and confronted with observations of nursing staff)<br>-S1: Patients motivational level throughout the study (from study introduction to end of patient participation).<br>-S2: Alliance to care during the 8 months of follow-up<br>-S3: Calculation of "cost-utility" and "cost-effectiveness"<br>-S4: Estimation of avoided costs and calculation of the differential with the actual consolidated cost of the NEHOTEL® program |
| Secondary outcome includes different objectives such as:<br>-“Improving the patients’ quality of life” measured by QLQ-30 and QLQ&HN-35 scores / number of therapeutic events completed compared to the initial objective / adverse events / quantified evolution of cancer biomarkers)<br>-“Evaluating the evolution undernutrition markers and the impact on the vital prognosis at 8 months” (biological markers of malnutrition during medical visits / food intake reported by the patient / consumption of Oral Nutritional Supplements / prognostic nutritional index PNI at 8 months)<br>-“Evaluating the effectiveness of non-dietetics reinforcement tools” (adapted physical activity-related BORG (rating perceived exertion) and IPAQ (International Physical Activity Questionnaire) scales / socio-aesthetic-related “DRAWING TEST” and ROSENBERG self-esteem scales / sophrology-related HAD (hospital anxiety and depression) and EQ-5D-VAS (health representation) scales during NEHOTEL® as well as the 8 months of follow-up)<br>-“Evaluating the effectiveness for the loved ones” (burden assessment Zarit scale) |

## Results

### Baseline concept

Because of the stress caused by the announcement of the disease and the amount of information to be integrated, the expert patients suggested that the supportive care program be built in the form of an immersive course on several consecutive days. A 5-days session appears to be the most suitable for a multidisciplinary team of qualified supportive care professionals to take turns, and involve the presence of a family member if possible. Such an initiative must be based on the understanding that patients need to take a break to integrate the dynamics of the new life awaiting them. The supporting care program is organized as a therapeutic education program (TEP) starting with an educational assessment of the in-patient and ending with an assessment of the skills acquired by the out-patient in response to the individual objectives of the educative contract. NEHOTEL^®^ approach is centered on dietetics and culinary skills, supported by incentive tools such as socio-aesthetics, adapted physical activity and sophrology. The educational objective is to promote empowerment and early autonomy in view to contain or reverse the spiral that increases the risks of undernutrition and morbimortality (Figure 1). It was suggested to gather 4 to 8 patients per session to create positive group dynamics. The first proposal was to integrate patients strictly just after the disease announcement and before the beginning of the treatment protocol. This point has been largely discussed with the medical staff and finally revised (see “discussion part”).

### Immersive integrated program

Careful step-by-step work with the team of supportive care professionals resulted in an education program, the synopsis of which is presented in Figure 2. The day of arrival is devoted to completing the information from the shared education assessment carried out beforehand at the addressing hospital, to design the educative contract for each patient, and to reinforce the commitments of the program (beliefs about the concept of health and education, quality of life, understanding of the illness, learning capacities, visualization of the benefits of NEHOTEL^®^, etc.). The second day focusses on potential occurrence of symptoms linked to the pathological situation and the side effects of the treatments. Nutritional and culinary advices, cooking workshops then follow through to provide reassuring solutions. Sophrology, socio-aesthetic care and adapted physical activity workshops are interspersed throughout the week in order to de-stress the group for a better cognitive ability. The last day is dedicated to the evaluation of the educational assessment of the participants leaving the center, with TEP professional and psychologist. Mid-week, a loved one is invited to stay for two nights on site, and activities for them are integrated into the program. If loved ones cannot come on site, an internet connection can be set up to work at particular moments. Finally, a patients’ association comes to meet the participants on the Wednesday evening to discuss with the group. Two sentinels, who are professional caregivers, take turns night and day, listening to participants and preparing the standard evening meals not included in the culinary sessions.

**Figure 2.**
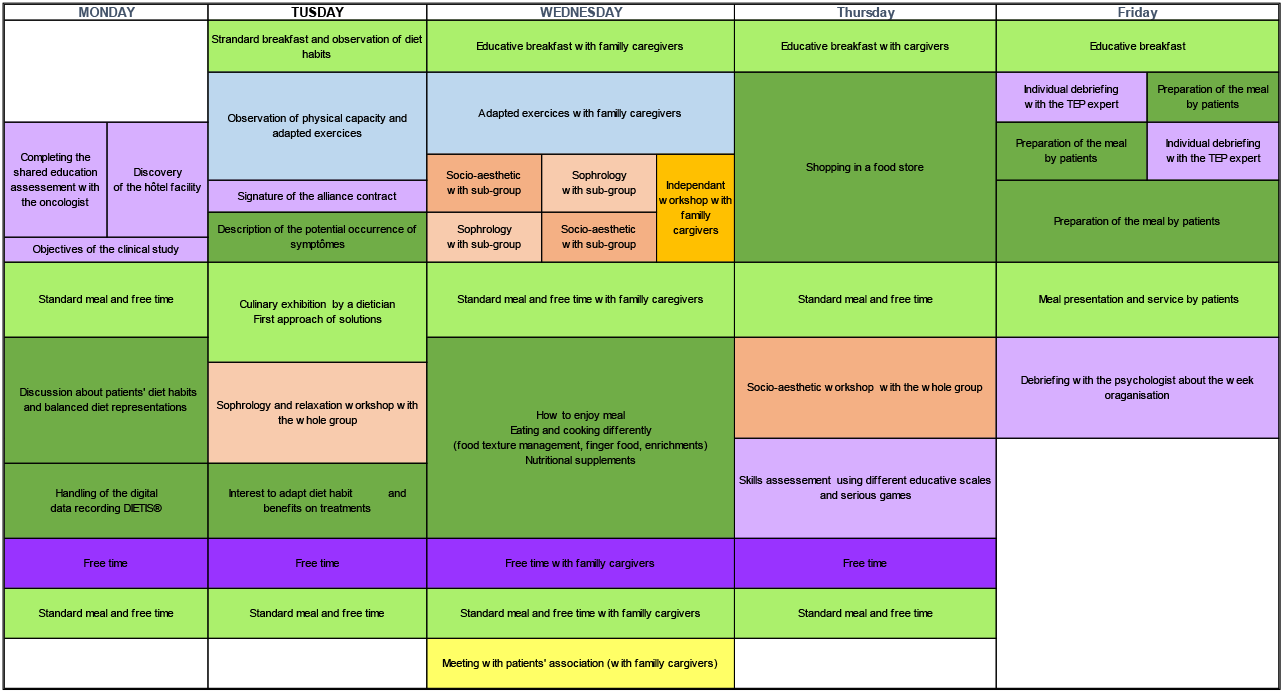
Synopsis of the 5-days supportive care program co-constructed by expert patients and supportive care professionals. Color coding: lavender: TEP meetings and assessments; light green: mealtime; green; culinary or nutritional workshops; purple: Free time; blue: physical activity; light salmon: sophrology; salmon: socio-aesthetic; gold: workshop specific for family caregiver; yellow: meeting with patients’ association.

### NEHOTEL Resort^®^ facilities proposed by expert patients

It was important for the expert patients that the reception site should be non-medical but located near a hospital, in a quiet place equipped and functioning like a hotel-restaurant offering an all-inclusive service. The venue is composed of individual rooms that can be shared during the visit of the family loved ones if they wish. The lounge, tea room and library allow for relaxation between workshops and in the evening. Evening meals are prepared on site and shared in the dining room. Both lunchtime cooking demonstrations as well as educational breakfasts are provided in a specific area by the dietary team. The socio-aesthetics room includes skin care products and massage chairs with selected musical, luminous and odorous atmosphere. The sophrology room is located in a quiet area equipped with a vegetal wall. A room and an outdoor area allow for adapted physical activities. The site has a reception desk and security surveillance at night. A minibus takes participants to do the educative shopping workshop in the city. The stay of the patient and the family caregiver is free of charge and the transportation of the patients is covered by prescription from the addressing practitioner.

## Discussion

“Is a nutrition and multimodal prehabilitation before cancer treatment conceivable as supportive care?” is the question addressed by expert patients to the medical staff. That question involves multiple medical, ethical and technico-economic considerations.

### An innovative hospitality facility

This hotel-restaurant facility equipped to receive newly diagnosed cancer patients does not exist in France. Some selected hotels close to hospitals are open to receive patients between medical visits or after hospital intervention, driven by an economic objective. Some spa resorts offer targeted nutritional support care for women who have completed their hormone-dependent cancer treatment with a risk of weight gain. Some associations organize one-week stays with small groups of cured or remission patients. In this case, accommodation and meeting rooms are rented for the occasion in a holiday region (mountain, seaside) and the association organizes multidisciplinary supportive care sessions including psychology, socio-aesthetics and adapted physical activity. In contrast, the NEHOTEL Resort^®^ concept has a permanent staff working in a building strictly dedicated to this activity. Participants are addressed by the surrounding hospitals within a 2-hours transport range. The ideal would be to have about 10 permanent and standardized sites in France to cover the territory. The NEHOTEL Resort^®^ concept co-created with the expert patients and supportive care professionals has been considered functional by the medical staff. A pilot site has been set up within 5min to the local hospital. Because the NEHOTEL^®^ 5-days educative synopsis as well as the NEHOTEL Resort^®^ facility are considered innovative, intellectual property protection were sought for the concept and logos.

### Risk assessment of patient refusal to participate and/or medical organization limits

Proposing NEHOTEL^®^ before any treatment has been an important debate taking in account both patient acceptance and potential medical organization limits. In the NEODIA 2010-survey, 68% of respondents agreed that food could have an effect on the body during treatment [12]. To the question: “Would you have liked to have received nutritional or culinary information earlier in your care pathway to be more autonomous for meals?” the answer was positive at 69.5% (N=137/197), of which 91.2% (N=125/137) were women. To the question: “Would you come to a nutrition supportive care workshop before treatment?”, 64.2% (N=88/137) of these patients think they would “accept” (N=26/137) or “definitely accept” (N=62/137), while 35.8% (N=49/137) would “definitely refuse”. These results highlighted that nutrition supportive care programs throughout the medical care protocols, whether in hospital settings or in city setting via patients’ associations, as it currently exists in France, fully have their place. However, these and other data also show that a significant percentage of people were seeking early information, even agreeing to attend a workshop before treatment [22]. Unsurprisingly, these were mostly women, who buy food for the family and are implicated in cooking. It is estimated that supportive care actions in oncology only reach about 30 to 40% of patients in our territory of Hauts-de-France, favoring urban populations (Onco Hauts-de-France network, personal communication). Both the accessibility and the disparities in the distribution of supportive care could be an explanation. In case of undernutrition that have been diagnosed, far too many patients are addressed to dieticians at a late stage. They then find it difficult to apply these specific dietary practices to tackle undernutrition [24, 25], thus reducing their motivation and compliance. In the NEODIA survey it appears that a significant percentage of patients prefer to wait for adverse events to occur before making any changes. However, it remains important to propose a prehabilitation course allowing to record a lot of information beforehand through a multidisciplinary approach, relayed by the usual supportive care schedule in nutrition alongside the therapeutic care protocol.

The role and scope of nutritional prehabilitation in cancer management is becoming increasingly accepted [23, 26]. However, not all oncology departments have a pre-discharge consultation, as is the case in Switzerland or Belgium. This could be detrimental to recruitment in the NEHOTEL^®^ study. Good coordination of the professionals involved after the announcement of the disease will be essential to integrate NEHOTEL^®^ without necessarily delaying the start of treatment. Particularly in HN cancer situations, malnutrition affects the clinical decision to resect the tumor, which is the main and potentially curative step in the management of this type of cancer [17, 25]. It is why the medical staff of NEHOTEL^®^ will also include patients after preventive surgery for clinical nutrition (veinous access, gastrostomy) and up to the second chemotherapy/radiotherapy cure and not only patients just after the disease announcement, as initially suggested by the expert patients. We believe that this decision will relieve some of the concerns addressed by oncology teams while still being useful as a prehabilitation tool for the patients.

## Conclusion

Supporting care before, during and after treatments should be integrated throughout the oncology continuum and delivered by trained professionals with the expertise to diagnose and treat a patient’s nutritional, physical, psychological and cognitive impairments. Prehabilitation as soon as possible presents an opportunity to improve patient’s quality of life and mental well-being in preparation for intensive treatments involving surgery, chemotherapy, and/or radiotherapy. Benefits include empowerment, improved functional independence, reduced comorbidity and associated treatments, and also reduced health care expenditures. The multidisciplinary medical staff need to consider the nutritional status of cancer patients before they define a treatment schedule. It would be unethical to treat undernourished patients as it increases risks of treatment severity or toxicity: the right patient, the right care, at the right time.

The identification and management of unmet supportive care needs in nutrition are an essential component of cancer health care in order to guide the development of new services to improve quality of life and prognostic. Research in nutrition (what foods, accepted supplements, when to eat, how much, culinary preparation) are more operational when patients are included in multidisciplinary scientific teams. Input is necessary from patients, their loved ones, oncologists, dietitians and nutrition experts, occupational therapists, and persons versed in cross-cultural issues on nutrition such as catering. The expanding role for nutrition gives us one more tool for supportive care in oncology. The WHO has stated that improved adherence to the treatment of chronic diseases, of which cancer is one, would have more benefit than any potential medical discovery. Nutrition prehabilitation may enhance quality of life, chance to complete treatments, therapeutic benefits, and improve recovery patterns. This holistic approach to patients’ demands must nevertheless be framed by evidence-based clinical research.

## Data Availability

All data produced in the present study are available upon reasonable request to the authors (study protocol not started)

## Abbreviations

AFSOS: association francophone de soins oncologiques de support
ARS: Agence régionale de santé
BMI: body mass index
CNCR: conseil national de coordination de la recherche
CONSORT: consolidated standards of reporting trials
DRESS: direction de la recherche, de l’enseignement supérieur et des formations sanitaires et sociales
FHF: fédération hospitalière de France
HAD: hospital anxiety and depression; HN: head and neck
I: inclusion
IDDSI: international dysphagia diet standardization initiative
IPAQ: international physical activity questionnaire
MSG: monosodium glutamate
NI: non inclusion
NPIS: non-pharmaceutical intervention solution
P: primary; PNA: programme national alimentation
PNI: prognostic nutritional index
QLQ: quality of life questionnaire
S: secondary
SIRHA: salon international de la restauration hôtellerie et alimentation
SIRS: systemic inflammatory response syndrome
TEP: therapeutic education of patient
TNM: tumor node metastasis
VAS: visual/verbal analogue scale
WHO: world health organisation

## Acknowledgements

The translational research global program NEODIA is labeled and funded by the regional health agency (ARS) of Hauts-de-France, the French National Program of Diet (PNA), and the French league against cancer (departmental committee of Oise). NEODIA was awarded by the 2017 World Hospitality and Food Service Event (SIRHA) and the AFSOS 2017 supportive care project in oncology. The NEHOTEL® concept emerging from NEODIA in 2019 was labeled by the Hospital French Federation (FHF) and the National Coordination Committee of Research (CNCR) as Excellence Research Territorial Program 2019, and by the GALIEN Price Official Selection 2020, 2021.

Approval for NEHOTEL® clinical study (EUDRACT 2021-A00901-40) was granted by the Ethics Committee of “Ile de France III” on July the 6th 2022 under the reference 21.04970.003904. International ClinicalTrials.gov identifier is NCT05495165. The clinical study is co-funded by the Research Program 2021 of the Hauts-De-France Regional Council delivered by the “Direction de la recherche, de l’enseignement supérieur et des formations sanitaires et sociales” (DRESS), and by the Malakoff Humanis group.

We would like to thank Dr François Browet (MD, Department of Gastroenterology and Surgery, Simone Veil Medical Center) and Dr Sif Bendjaballah (MD, Department of anatomo-pathology, Simone Veil Medical Center) for their contribution to the initiation of the NEHOTEL^®^ clinical protocol. We would also like to particularly thank the expert patient’s panel of the NEODIA translational research program as well as three UniLaSalle Engineers trainees, Emma Lopez, Clara Rousseau and Solange De Coudenhove, involved in the project initiation.

## Conflicts of interests

All authors certify that they have no affiliations with or involvement in any organization or entity with any financial interest or non-financial interest in the subject matter or materials discussed in this manuscript.

## Notes

### Competing Interest Statement

The authors have declared no competing interest.

### Clinical Trial

NCT05495165

### Author Declarations

Ethics Committee of Ile de France III gave ethical approval for this work on July the 6th 2022 under the reference 21.04970.003904. International ClinicalTrials.gov identifier is NCT05495165.

